# Quantification of the spread of SARS-CoV-2 variant B.1.1.7 in Switzerland

**DOI:** 10.1101/2021.03.05.21252520

**Authors:** Chaoran Chen, Sarah Nadeau, Ivan Topolsky, Marc Manceau, Jana S. Huisman, Kim Philipp Jablonski, Lara Fuhrmann, David Dreifuss, Katharina Jahn, Christiane Beckmann, Maurice Redondo, Christoph Noppen, Lorenz Risch, Martin Risch, Nadia Wohlwend, Sinem Kas, Thomas Bodmer, Tim Roloff, Madlen Stange, Adrian Egli, Isabella Eckerle, Laurent Kaiser, Rebecca Denes, Mirjam Feldkamp, Ina Nissen, Natascha Santacroce, Elodie Burcklen, Catharine Aquino, Andreia Cabral de Gouvea, Maria Domenica Moccia, Simon Grüter, Timothy Sykes, Lennart Opitz, Griffin White, Laura Neff, Doris Popovic, Andrea Patrignani, Jay Tracy, Ralph Schlapbach, Emmanouil T. Dermitzakis, Keith Harshman, Ioannis Xenarios, Henri Pegeot, Lorenzo Cerutti, Deborah Penet, Anthony Blin, Melyssa Elies, Christian L. Althaus, Christian Beisel, Niko Beerenwinkel, Martin Ackermann, Tanja Stadler

**Affiliations:** Department of Biosystems Science and Engineering, ETH Zürich, Basel, Switzerland; Swiss Institute of Bioinformatics, Switzerland; Department of Environmental Systems Science, ETH Zürich, Swiss Federal Institute of Technology, Zurich, Switzerland; Viollier AG, Allschwil, Switzerland; Dr Risch, Labormedizinisches Zentrum, Switzerland; Clinical Bacteriology and Mycology, University Hospital Basel, Basel, Switzerland; Applied Microbiology Research, Department of Biomedicine, University of Basel, Basel, Switzerland; Geneva Center for Emerging Viral Diseases and Laboratory of Virology, Geneva University Hospitals, Geneva, Switzerland; Genomic Facility Basel, Department of Biosystems Science and Engineering, ETH Zürich, Basel, Switzerland; Functional Genomics Center Zurich, ETH Zürich and University of Zurich, Zurich, Switzerland; Health 2030 Genome Center, Geneva, Switzerland; University of Geneva Medical School, Geneva, Switzerland; Institute of Social and Preventive Medicine, University of Bern, Bern, Switzerland; Department of Environmental Microbiology, Eawag, Dubendorf, Switzerland; Department of Microbiology and Molecular Medicine, Faculty of Medicine, University of Geneva, Geneva, Switzerland; Division of Infectious Diseases, Geneva University Hospitals, Geneva, Switzerland; Department of Medicine, Faculty of Medicine, University of Geneva, Geneva, Switzerland

**Keywords:** Pandemic, SARS-CoV-2, COVID-19, B.1.1.7, transmission advantage

## Abstract

**Background:** In December 2020, the United Kingdom (UK) reported a SARS-CoV-2 Variant of Concern (VoC) which is now named B.1.1.7. Based on initial data from the UK and later data from other countries, this variant was estimated to have a transmission fitness advantage of around 40-80% [1, 2, 3].

**Aim:** This study aims to estimate the transmission fitness advantage and the effective reproductive number of B.1.1.7 through time based on data from Switzerland.

**Methods:** We generated whole genome sequences from 11.8% of all confirmed SARS-CoV-2 cases in Switzerland between 14 December 2020 and 11 March 2021. Based on these data, we determine the daily frequency of the B.1.1.7 variant and quantify the variant’s transmission fitness advantage on a national and a regional scale.

**Results:** We estimate B.1.1.7 had a transmission fitness advantage of 43-52% compared to the other variants circulating in Switzerland during the study period. Further, we estimate B.1.1.7 had a reproductive number above 1 from 01 January 2021 until the end of the study period, compared to below 1 for the other variants. Specifically, we estimate the reproductive number for B.1.1.7 was 1.24 [1.07-1.41] from 01 January until 17 January 2021 and 1.18 [1.06-1.30] from 18 January until 01 March 2021 based on the whole genome sequencing data. From 10 March to 16 March 2021, once B.1.1.7 was dominant, we estimate the reproductive number was 1.14 [1.00-1.26] based on all confirmed cases. For reference, Switzerland applied more non-pharmaceutical interventions to combat SARS-CoV-2 on 18 January 2021 and lifted some measures again on 01 March 2021.

**Conclusion:** The observed increase in B.1.1.7 frequency in Switzerland during the study period is as expected based on observations in the UK. In absolute numbers, B.1.1.7 increased exponentially with an estimated doubling time of around 2-3.5 weeks. To monitor the ongoing spread of B.1.1.7, our plots are available online.

## 1 Introduction

In mid-December 2020 a SARS-CoV-2 variant named B.1.1.7 was first reported as more transmissible than previously circulating strains [4, 5, 6]. This variant, whose name comes from the pangolin lineage nomenclature [7] and which was first identified in the UK, carries the N501Y mutation in the spike protein which may increase ACE2 receptor affinity [8]. Within only a few months, B.1.1.7 became the dominant variant in the UK epidemic.

Based on these first reports, Switzerland began an intense effort to detect and trace B.1.1.7 [9]. The first infections with B.1.1.7 in Switzerland were confirmed on 24 December 2020 and retrospective analyses identified B.1.1.7 in samples dating back to October [9]. In total, 1370 infections with B.1.1.7 were confirmed up to 05 February 2021 [9].

Observing a variant increase in frequency does not necessarily mean it has a transmission fitness advantage. For example, a variant named 20A.EU1 spread rapidly across Europe in summer 2020. However, data suggests that extended travel and superspreading events caused that spread, not a viral transmission fitness advantage [10]. Alternatively, partial immune escape may help a variant spread compared to other variants.

In the case of B.1.1.7, Davies et al. [3] concluded that the variant’s spread is poorly explained by a hypothesis of immune escape. Instead, B.1.1.7’s rapid increase in frequency in many high-prevalence regions across the UK in parallel is well-explained by a transmission fitness advantage [3]. Indeed, several different analyses based on UK data suggest B.1.1.7 has a transmission fitness advantage between 40 and 80% [1, 2, 3]. Davies et al. [3] also obtain similar estimates based on data from Denmark. Finally, analysis of the spread of the N501Y mutation in Switzerland suggests a similar transmission fitness advantage [11].

In this study, we determine the frequency of B.1.1.7 through time in Switzerland and calculate its transmission fitness advantage based on three different datasets. First, we generated whole-genome sequences from randomly selected samples provided by the diagnostics company Viollier AG. Second, we use data from the diagnostics company Dr Risch AG which screens all their samples for B.1.1.7. Finally, we use whole-genome sequences generated from all patients who tested positive for SARS-CoV-2 at the University Hospital Geneva (HUG) with CT values below 32 beginning 23 December 2020. These three datasets are differently representative of Switzerland at the national and regional levels. Based on these data, we quantified the transmission fitness advantage of B.1.1.7 for Switzerland as well as for the seven Swiss economic regions (Grossregionen). We additionally calculated the reproductive number for B.1.1.7 and show how the number of B.1.1.7 infections developed through time.

The core plots presented here were regularly updated between mid-January and mid-April 2021 on[12] and, since then are available on the CoV-Spectrum website [13]. Some of the results are additionally displayed on the Swiss National COVID-19 Science Task Force website as of May 2021 [14]. The code and data used for this study are publicly available on Github [15]. All sequences are available on GISAID [16] (section A.7).

## 2 Methods

### 2.1 Data

We analyze three different datasets. The primary dataset is a set of whole-genome sequences generated from samples provided by Viollier AG, a large Swiss diagnostics company that processes SARS-CoV-2 samples from across Switzerland. Each week, a random selection of samples from amongst all positive tests processed by the company were selected for whole-genome sequencing. Each sample is associated with a test date and the canton in which the test was performed. Whole-genome sequences were generated from selected samples according to the procedures described in the supplementary materials section A.1. We define a sequence to be a B.1.1.7 sample if at least 80% of the lineage-defining, non-synonymous nucleotide changes according to [4] are present.

The second dataset is daily counts of B.1.1.7 infections amongst tests processed by Dr Risch AG, another Swiss diagnostics company. Each count is associated with a test date but not a geographic location. The screening procedures for B.1.1.7 used to generate these data are described in the supplementary materials section A.2.

The third and final dataset is a set of whole-genome sequences generated from patients at the University Hospital Geneva (HUG). Samples from all patients who tested positive there with a CT of below 32 were sent for whole-genome sequencing, which were generated according to the procedures described in the supplementary materials section A.1. As with the first dataset, a sequence is defined to be a B.1.1.7 sample if at least 80% of the lineage-defining, non-synonymous nucleotide changes are present.

These three datasets differ in their size and geographic representation. The first dataset includes 9772 sequences from the study period 14 December 2020 to 11 March 2021 (approximately 780 per week, status current as of 23 March 2021). These data represent 5.3% of the 184,165 confirmed infections in Switzerland during this period. The second dataset includes 12,019 samples screened for B.1.1.7. Taken together, these two datasets represent 11.8% of all confirmed SARS-CoV-2 infections in Switzerland during our study period. Finally, the third dataset is specific to the Lake Geneva region. It covers most of the study period, from 23 December 2020 to 04 March 2021, and includes 2074 sequences which represent 7% of all confirmed infections in the Lake Geneva region during that period.

Regarding geographic location, the diagnostic companies Viollier AG and Dr Risch AG both processes samples from all over Switzerland but the intensity varies across regions. The set of sequenced samples inherits this uneven geographical distribution. For example, relative to the total number of confirmed infections, the Viollier AG dataset includes over eight times more sequenced infections from the region Nordwestschweiz than from the region Ticino (Table S1). The Dr Risch AG data, on the other hand, has much better coverage of Eastern Switzerland (Table S2). In summary, these two national-level datasets differ in their geographic biases.

In what follows, we analyze the frequency of identified B.1.1.7 samples per day in the three datasets and compare our results between them. Both national-level datasets (Viollier AG and Dr Risch AG) are used to generate estimates on the national level, and comparing between them shows the effect of different geographic biases. Since only the Viollier AG dataset is resolved at the regional level, all regional-level estimates are based on this dataset. Finally, the HUG dataset specific to the Lake Geneva region allows a more detailed view on B.1.1.7 spread in this region and a validation of results generated using the other two datasets.

### 2.2 Statistical inference

We fit a logistic model to the frequency of B.1.1.7 samples per day to estimate the logistic growth rate *a* and the sigmoid’s midpoint *t*_0_. From that, we derive an estimate of the transmission fitness advantage of B.1.1.7 under a continuous (*f*_*c*_) and a discrete (*f*_*d*_) model. Each model could plausibly describe the actual dynamics, so we present results from both for comparison. Further, we estimate the reproductive number R for the B.1.1.7 and non-B.1.1.7 infections. The mathematical derivations are described in the supplementary materials in the sections A.3 and A.4. Finally, we show the projected number of confirmed infections in the future under the continuous model. We initialize the model on 01 January 2021 with the estimated number of B.1.1.7 and non-B.1.1.7 confirmed infections on that day. We assume a reproductive number for the non-B.1.1.7 infections as estimated on the national level for 01 January-17 January 2021. Further, we assume that the expected generation time is 4.8 days and the fitness advantage is the estimated *f*_*c*_ for the region and dataset of interest (Table 1).

**Table 1:**
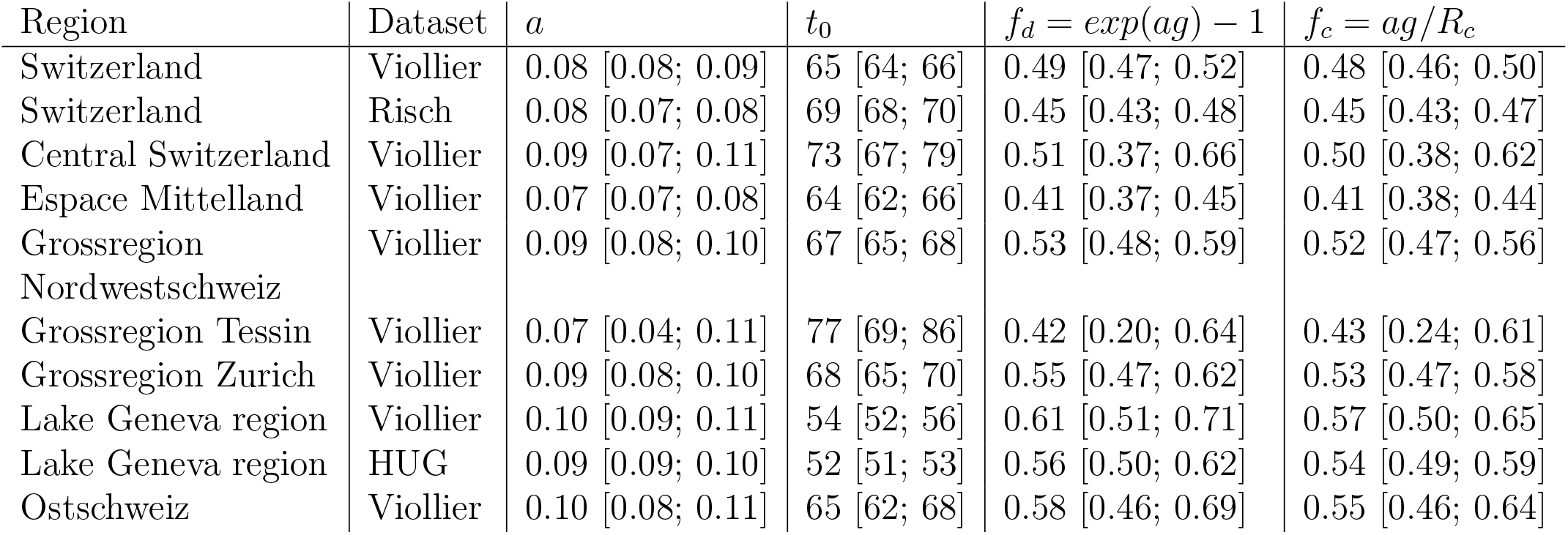
Estimates of the growth rate *a* and the sigmoid’s midpoint *t*_0_ (measured in days after Dec. 14)as well as the transmission fitness advantages *f*_*d*_ and *f*_*c*_ are reported. In the *f*_*c*_ calculation, the Swiss-wide estimate of the reproductive number for the time period 01 January 2021-17 January 2021 is assumed for the *R*_*c*_.

| Region | Dataset | $a$ | $t_0$ | $f_d = \exp(ag) - 1$ | $f_c = ag/R_c$ |
| --- | --- | --- | --- | --- | --- |
| Switzerland | Viollier | 0.08 [0.08; 0.09] | 65 [64; 66] | 0.49 [0.47; 0.52] | 0.48 [0.46; 0.50] |
| Switzerland | Risch | 0.08 [0.07; 0.08] | 69 [68; 70] | 0.45 [0.43; 0.48] | 0.45 [0.43; 0.47] |
| Central Switzerland | Viollier | 0.09 [0.07; 0.11] | 73 [67; 79] | 0.51 [0.37; 0.66] | 0.50 [0.38; 0.62] |
| Espace Mittelland | Viollier | 0.07 [0.07; 0.08] | 64 [62; 66] | 0.41 [0.37; 0.45] | 0.41 [0.38; 0.44] |
| Grossregion Nordwestschweiz | Viollier | 0.09 [0.08; 0.10] | 67 [65; 68] | 0.53 [0.48; 0.59] | 0.52 [0.47; 0.56] |
| Grossregion Tessin | Viollier | 0.07 [0.04; 0.11] | 77 [69; 86] | 0.42 [0.20; 0.64] | 0.43 [0.24; 0.61] |
| Grossregion Zurich | Viollier | 0.09 [0.08; 0.10] | 68 [65; 70] | 0.55 [0.47; 0.62] | 0.53 [0.47; 0.58] |
| Lake Geneva region | Viollier | 0.10 [0.09; 0.11] | 54 [52; 56] | 0.61 [0.51; 0.71] | 0.57 [0.50; 0.65] |
| Lake Geneva region | HUG | 0.09 [0.09; 0.10] | 52 [51; 53] | 0.56 [0.50; 0.62] | 0.54 [0.49; 0.59] |
| Ostschweiz | Viollier | 0.10 [0.08; 0.11] | 65 [62; 68] | 0.58 [0.46; 0.69] | 0.55 [0.46; 0.64] |

## 3 Results

We estimate the logistic growth rate *a* and the sigmoid’s midpoint *t*_0_ based on the two national-level datasets from Viollier AG and Dr Risch AG (Table 1). Taking the estimates of both datasets together, we obtain a growth rate *a* of 0.07-0.09 per day for Switzerland. For each economic region, the estimated uncertainty interval of a overlaps with the Swiss-wide uncertainty. We have little data for two out of seven regions (Ticino and Central Switzerland; *<*1100 sequences in total) resulting in very wide uncertainty intervals. From the *t*_0_ estimates, we observe that the Lake Geneva region was about 2 weeks ahead of the rest of Switzerland with respect to B.1.1.7 spread. This confirms estimates from [11]. Our initial analyses of the data in January 2021 projected that B.1.1.7 will become dominant in Switzerland in March 2021. Indeed, our latest data points suggest a frequency of B.1.1.7 in confirmed infections of around 80% for 11 March 2021.

In Fig. 1 and 2, we graphically illustrate the logistic growth in frequency of B.1.1.7 and show the daily data together with an estimate of the proportion of B.1.1.7 under the logistic growth model.

**Figure 1:**
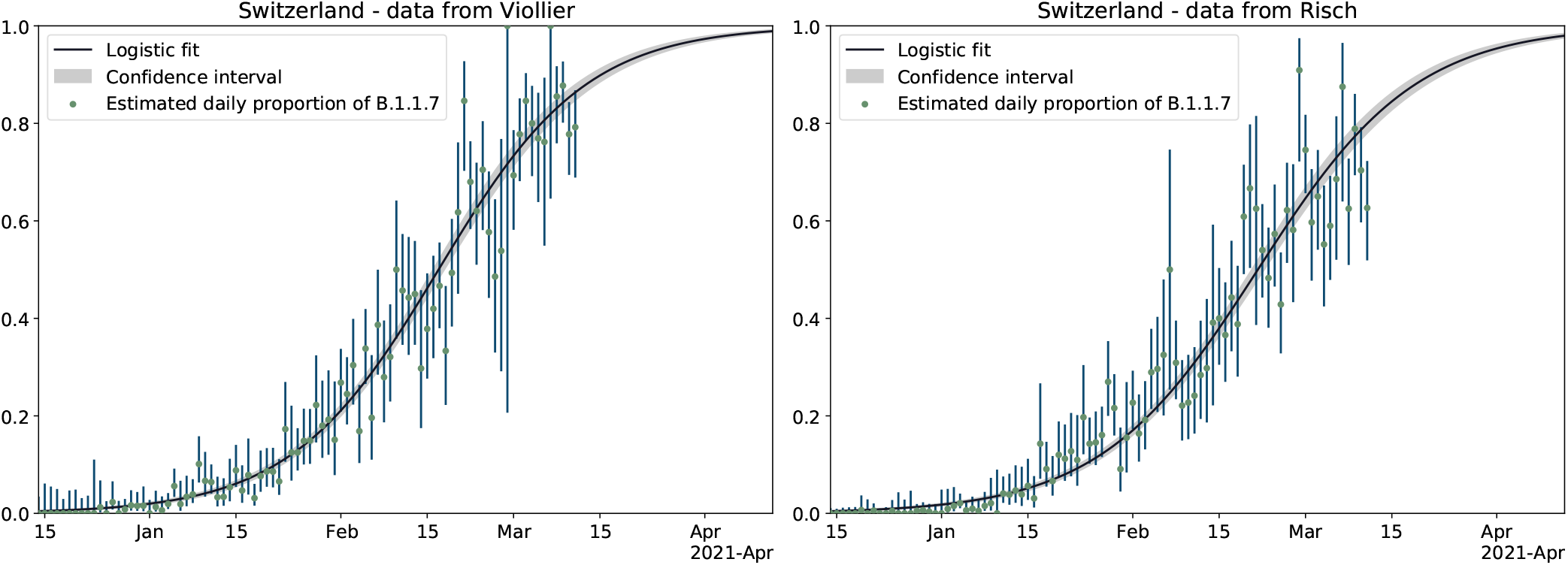
Logistic growth of frequency of B.1.1.7 in Switzerland. Green points are the empirical proportions of B.1.1.7 for each day (i.e. number of B.1.1.7 samples divided by the total number of samples). Blue vertical lines are the estimated 95 % uncertainty of this proportion for each day, assuming a simple binomial sampling and Wilson uncertainty intervals. A logistic growth function fit to the data from all of Switzerland is shown in black with the 95 % uncertainty interval of the proportions in gray (i.e. *p*(*t*) from Eqn. 2 and 5 in the supplementary material section A.3).

**Figure 2:**
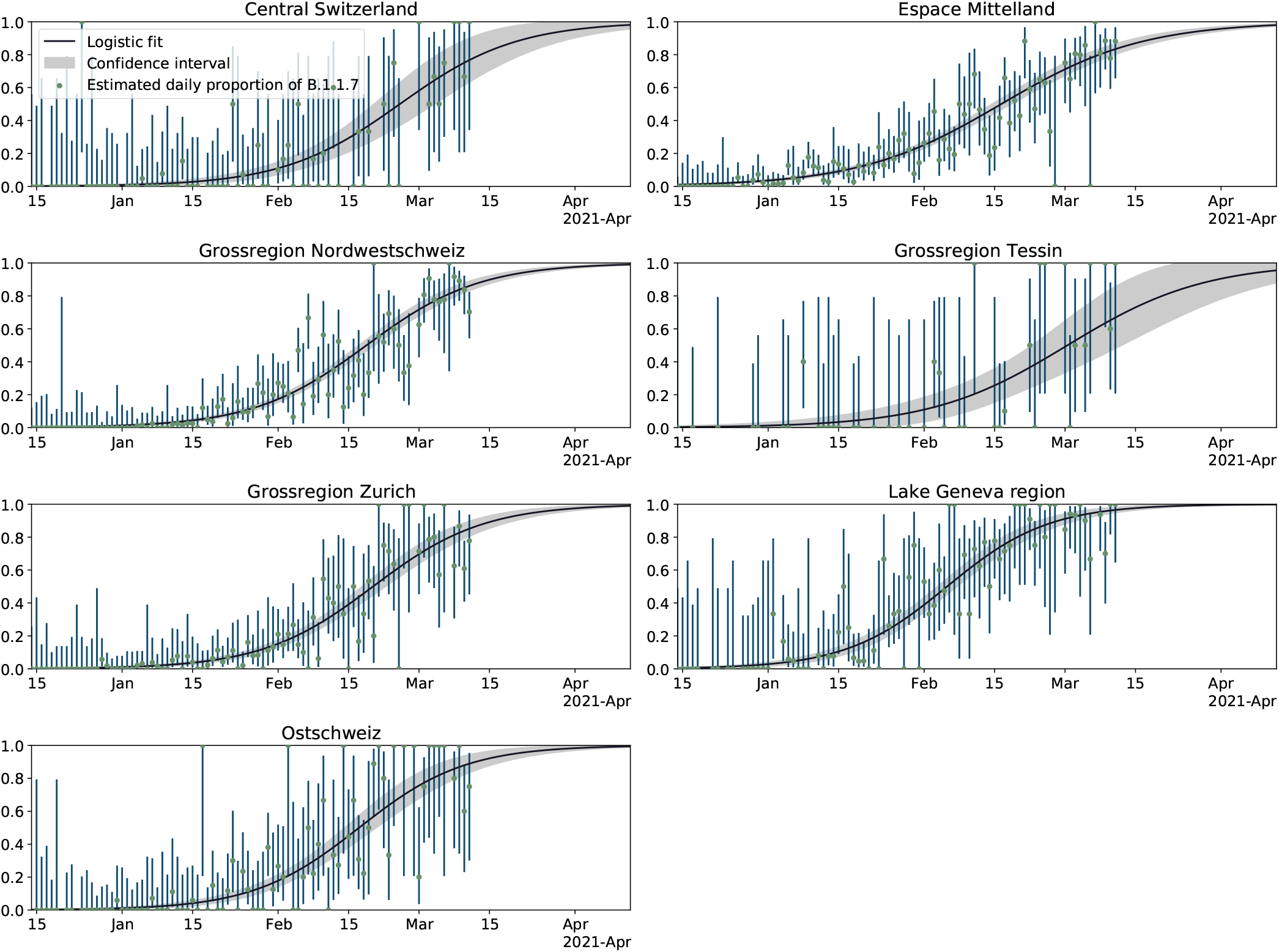
Logistic growth of frequency of B.1.1.7 in the seven economic regions of Switzerland. For details see legend of Fig. 1.

As a validation of the logistic growth parameter estimates, we additionally analyzed the third, Lake Geneva-specific dataset from HUG. The estimates for the Lake Geneva region based on Viollier AG data agree very well with these independent estimates based on HUG data (Table 1 and Figure S2).

Next, we estimate the reproductive number for B.1.1.7 and non-B.1.1.7 infections on a national scale (Fig. 3). We note that non-pharmaceutical interventions to combat SARS-CoV-2 spread in Switzerland were strengthened on 18 January 2021 and then relaxed on 01 March 2021. Between 01 January and 17 January 2021, the reproductive number for B.1.1.7 was significantly above 1 (Viollier AG dataset: 1.24 [1.07-1.41], Dr Risch AG dataset: 1.46 [1.21-1.72]) while the reproductive number of non-B.1.1.7 was below 1 (Viollier AG dataset: 0.83 [0.65-1.00], Dr Risch AG dataset: 0.81 [0.67-0.96]). The reproductive number for B.1.1.7 calculated based on the Viollier AG dataset did not drastically change after 18 January 2021 (1.18 [1.06-1.30]) and agrees well with the estimates based on the Dr Risch AG dataset (1.15 [1.01,1.29]). The non-B.1.1.7 reproductive number also did not drastically change after this timepoint (Viollier AG dataset: 0.80 [0.68-0.91], Dr Risch AG dataset: 0.85 [0.76,0.93]).

**Figure 3:**
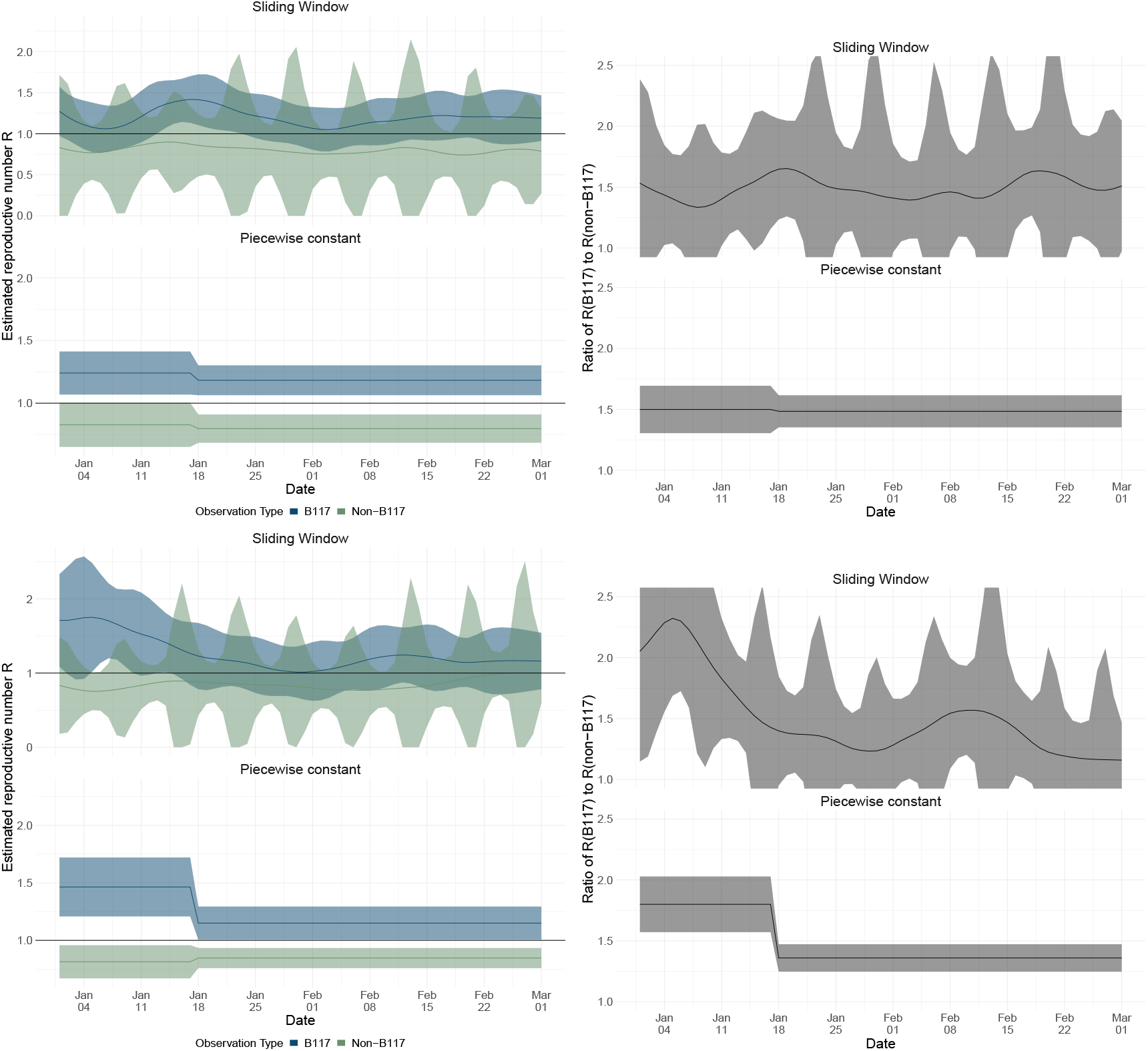
Estimates of the effective reproductive number *R* of the B.1.1.7 variant and non-B.1.1.7 variants. Results in the top row are based on Viollier data, and in the bottom row based on Risch data. Within each panel, the top row shows the results of the continuously varying R estimation, and the bottom of the piecewise constant *R* estimation. The left column shows the *R* estimates, whereas the right shows the ratio between *R* estimated for B.1.1.7 and *R* estimated for all non-B.1.1.7 variants. The confidence intervals for the *R* of non-B117 variants show a 7-day periodicity due to lower case reporting on weekends. The *R* value was allowed to change on 18 January 2021 in the statistical inference as measures were tightened on that day.

As expected from assuming a constant B.1.1.7 transmission fitness advantage, the ratio of the reproductive numbers for B.1.1.7 and non-B.1.1.7 based on the Viollier AG dataset was roughly constant throughout January and February. However, based on the Dr Risch AG dataset, the ratio of the reproductive numbers for B.1.1.7 and non-B.1.1.7 unexpectedly dropped in January. We suspect a potential bias in this dataset (such as preferential inclusion of B.1.1.7 in early January) because all estimates for B.1.1.7 in early January agree except those based on the Dr Risch AG dataset.

Our data does not allow us to estimate the variant-specific reproductive number for March yet. However, for the time period 10 March-16 March 2021, we can estimate the reproductive number based on all confirmed infections [17] (1.14 [1.00-1.26]). Since this estimate is based on confirmed cases from the second part of March when we project around 90-95% of all confirmed infections are B.1.1.7, this estimate should only slightly underestimate the reproductive number of B.1.1.7. In summary, we estimate the reproductive number for B.1.1.7 was above 1 since January 2021 while the reproductive number for non-B.1.1.7 variants was below 1.

Next, we calculate the transmission fitness advantage *f*_*c*_ of B.1.1.7 under our continuous-time model using the average reproductive number estimated between 01 January 2021-17 January 2021. Further, we calculate the transmission fitness advantage *f*_*d*_ under a discrete-time model. In both cases, we assume a generation time *g* of 4.8 days (the same as the mean generation time used to estimate the reproductive number). Table 1 shows the estimated fitness values under both methods. On a national level, we estimate a fitness advantage of 43-52% across methods and datasets. The regional estimates overlap with this interval. We note that we use the national reproductive number for the regional *f*_*c*_ estimates. Since Ticino had a lower reproductive number averaged over all variants [17], the *f*_*c*_ for Ticino may be an underestimate. Similarly, the Lake Geneva region had a higher reproductive number so the *f*_*c*_ for Lake Geneva may be an overestimate.

Finally, we show the projected dynamics of the epidemic under the continuous model using parameter values based on epidemic conditions in the first half of January (Fig. 4 and 5). We show how the number of B.1.1.7 infections develops over time in blue and how the number of non-B.1.1.7 infections develops in green. In particular, in January 2021, the model projects a decline in overall infections due to a decline in non-B.1.1.7 variants. However, the number of B.1.1.7 variants increases. Under this model, the total number of infections will increase again once B.1.1.7 becomes dominant. It is important to note that this simple model is intended to highlight whether epidemic dynamics change compared to early January 2021 due to B.1.1.7, and thus it assumes the transmission dynamics are constant. In particular, the model does not include effects of non-pharmaceutical interventions introduced on 18 January 2021 or removed on 01 March 2021, vaccination, immunity after infection, and population heterogeneity.

**Figure 4:**
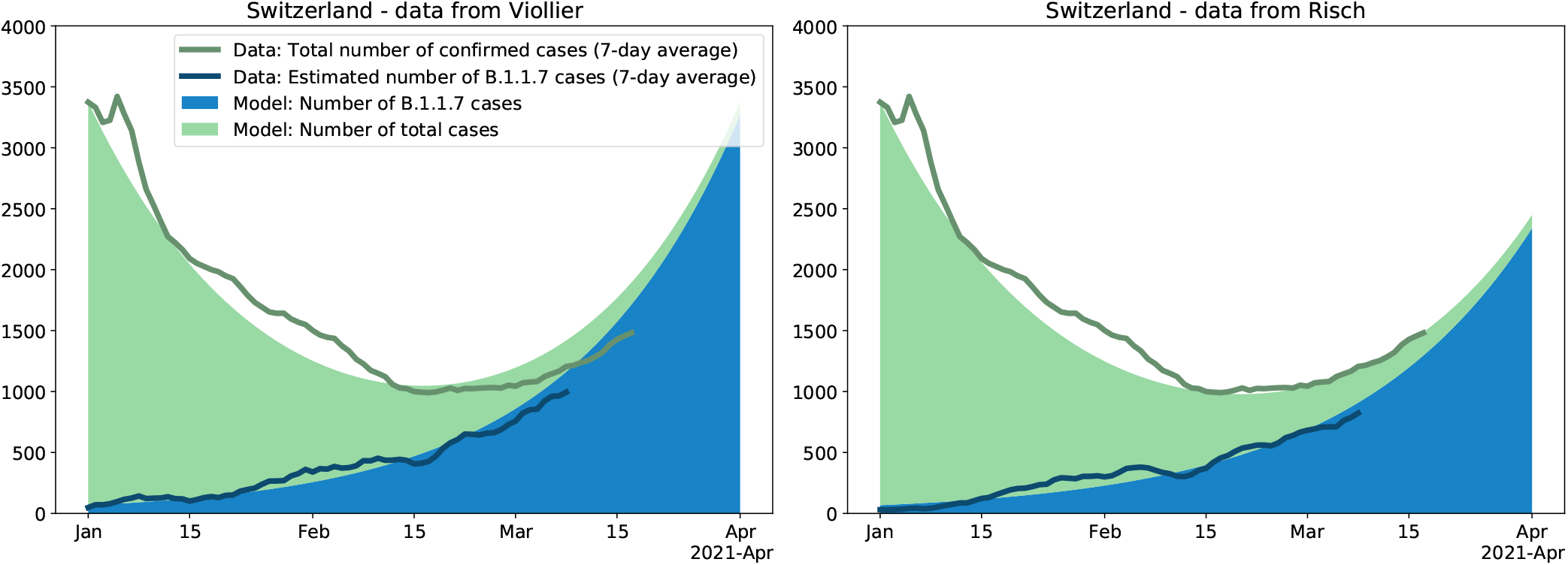
Change in the number of B.1.1.7 variants and in the number of all cases through time for Switzerland. Based on the average reproductive number *R*_*c*_ for Switzerland estimated for the time period 01 January 2021-17 January 2021 (i.e. prior to the tightening of measures on 18 January 2021) and the transmission fitness advantage *f*_*c*_ for the same time period, we plot the expected number of B.1.1.7 variants (blue) and the expected number of non-B.1.1.7 variants (green) under the continuous model. The model is initialized on Jan. 1 with the total number of cases and the estimated number of B.1.1.7 cases on that day. This model is compared to data: The dark green line is the total number of confirmed cases (7-day average). The dark blue line is the estimated number of confirmed B.1.1.7 cases (7-day average); this number is the product of the total number of confirmed cases for a day by the proportion of the B.1.1.7 variant for that day. If the empirical data develops as the model, the dark blue line follows the upper end of the blue area and the dark green line follows the upper end of the green area.

**Figure 5:**
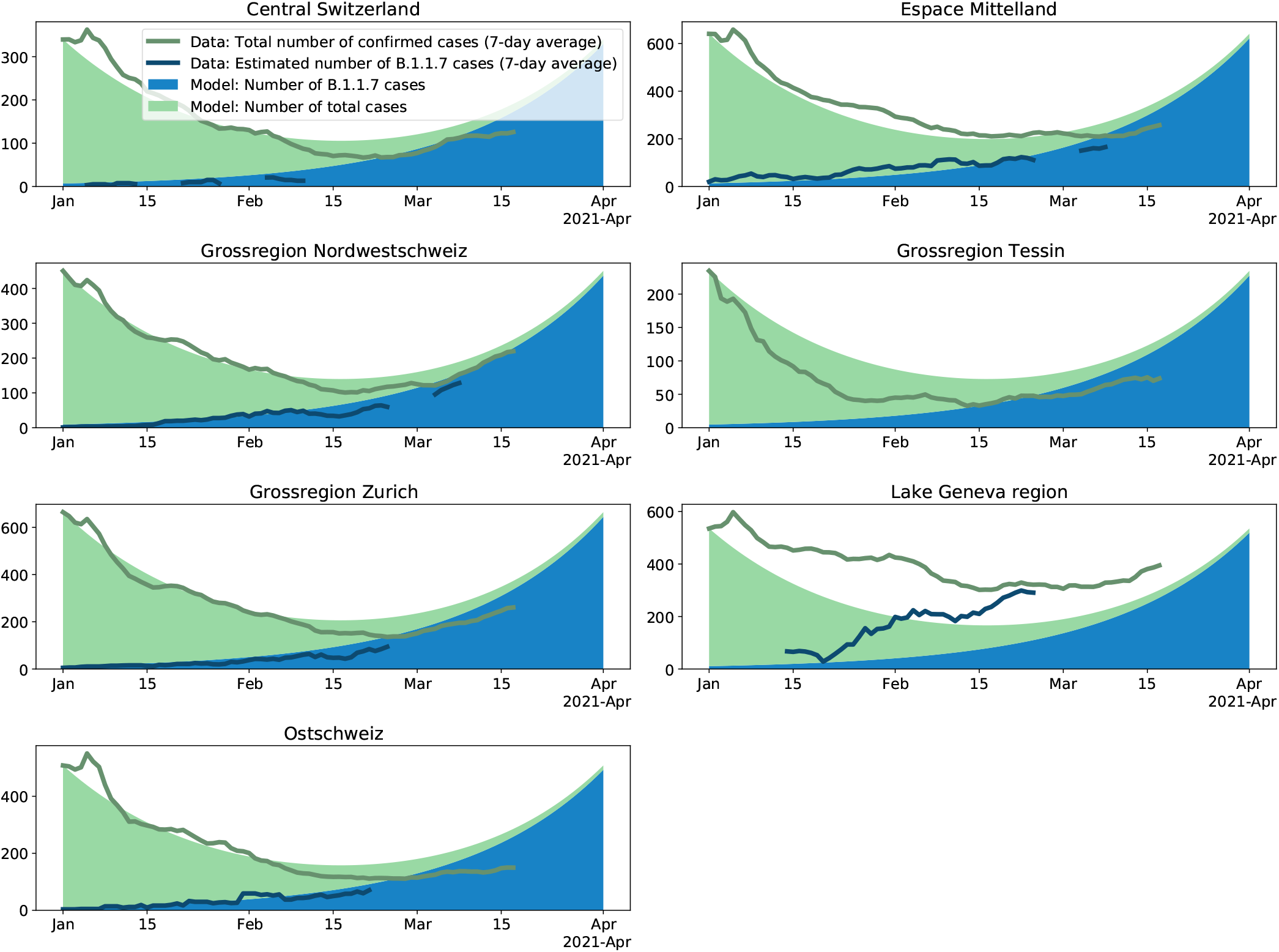
Change in the number of B.1.1.7 variants and in the number of all cases through time for the seven Swiss economic regions. For details see legend of Fig. 4. We use the reproductive number estimated for the whole of Switzerland for the continuous-time model such that we can compare to what extend regions differ from the national dynamic. The regional transmission fitness advantage is taken from Table 1.

In our projections, we assume that dynamics remained unchanged since early January. In order to explore to what extend this assumption is violated, we investigate whether empirical infection numbers follow the model-based projections. In Fig. 4 and 5, we show that the model (solid areas) and empirical data (lines) follow the same trajectories throughout January 2021 across datasets and regions, with the exception of the Ticino and Lake Geneva regions. As discussed above, we believe these discrepancies are a result of regional differences in the reproductive number (see also the supplementary materials, section A.5). After January 2021, we notice a discrepancy between the empirical data and the projections at the national level based on the Viollier AG dataset (Fig. 4) as well as the five regions with a good match in January (Fig. 5, also based on the Viollier AG dataset). The empirical data are below the model projections. This discrepancy is consistent with a slight reduction in the reproductive number after 18 January 2021.

On the other hand, the empirical data follow model projections using parameters estimated from the Dr Risch AG dataset very well until March (Fig. 4). Thus, changing the model from the Viollier AG-based parameters (R-value 0.83, fitness advantage 0.48) to the Dr Risch AG-based parameters (R-value 0.81, fitness advantage 0.45) leads to a very good fit until mid-March, highlighting that in a phase of overall exponential growth, slight changes of the parameters can have large consequences on the total number of infections. In other words, a slight reduction of transmission in late January 2021 and/or a slight misspecification of the Viollier AG-based parameters can explain the recent mismatch of the total number of infections and the Viollier AG-based projections.

## Discussion

We quantified the Swiss-wide transmission fitness advantage and the effective reproductive number of B.1.1.7 based on two national-level datasets. One dataset also allows us to obtain estimates specific to the seven Swiss economic regions. Swiss-wide estimates point towards a transmission fitness advantage of 43-52%. Based on our early sequencing data, we already projected in January 2021 that B.1.1.7 will become dominant in March. The same conclusion was reached by [11] in January 2021 based on Swiss data tracking S:501Y mutations. Indeed, on 11 March 2021 around 80% of characterized infections carried the B.1.1.7 variant. As of the date of writing, 26 March 2021, we expect almost all new infections may be caused by B.1.1.7 since cases are confirmed 8-11 days after the time of infection in Switzerland. Thus, the Swiss epidemic is now a B.1.1.7 epidemic.

The increase in the frequency of B.1.1.7 in Switzerland occurred as expected given the variant’s large transmission advantage. Unless measures such as contact tracing particularly target B.1.1.7, the variant’s transmission advantage causes it to dominate. However, the speed at which a variant increases in absolute numbers is a function of variables such as non-pharmaceutical interventions, adherence to such measures, and levels of immunity in the population.

We show that in the first half of January 2021, the absolute numbers of B.1.1.7 infections increased (R-value 1.24 [1.07-1.41] for 01 January-17 January2021 based on our Swiss-wide sequence dataset from Viollier AG) while the absolute numbers of all other infections decreased (R-value 0.83 [0.65-1.00]). For the second half of January 2021, the reproductive number for B.1.1.7 and non-B.1.1.7 infections only decreased slightly according to the Viollier AG data.

To facilitate a comparison of the B.1.1.7 reproductive number before and after the introduction of more non-pharmaceutical interventions on 18 January 2021, we note that the doubling time for a reproductive number of 1.24 is 15 days compared to 20 days for a reproductive number of 1.18, which makes a substantial difference in the overall epidemic dynamics.

Estimates based on the Swiss-wide Dr Risch AG data agree very well with this pattern, with the exception of B.1.1.7 in early January where a higher reproductive number is estimated. We speculate that the large drop in the reproductive number for B.1.1.7 based on the Dr Risch AG data is due to a bias in the early B.1.1.7 data, which possibly contained samples preferentially stemming from infections.

We estimate the reproductive number averaged over all confirmed infections in Switzerland from 10 March to 16 March 2121 (among which B.1.1.7 is projected to have caused 90-95% of infections) was 1.14 [1.00-1.26]. Taken together, these reproductive number estimates highlight that B.1.1.7 spread exponentially in Switzerland beginning in early January 2021. Our point estimate for the doubling time is around 2-3.5 weeks.

When comparing confirmed infections to model-based projections, we observe that the number of confirmed infections is lower in February 2021 than expected based on the model fit to Viollier AG data and using parameter estimates from the first half of January 2021. This may reflect a small overall slow-down in transmission, which is also suggested by our reproductive number estimates. However, this slow-down was not large and the reproductive number estimates are quite uncertain. On the other hand, the number of confirmed infections agrees very well with the model fit to Dr Risch AG data. We suggest that either a slight slow-down in transmission in late January and/or a slight misspecification of the Viollier AG-based model parameters may explain the discrepancy (assumed parameters are: non-B.1.1.7 reproductive number of 0.83 vs 0.81 and B.1.1.7 transmission fitness advantage of 0.48 vs 0.45 from Viollier AG and Dr Risch AG data, respectively). In summary, the reduction seen in the Viollier AG data may be the outcome of the strengthening of non-pharmaceutical interventions on 18 January or a slight bias in the parameter estimates from early January 2021. The 11.5% of confirmed infections characterized in our dataset is not sufficient to quantify the change in dynamics precisely enough to evaluate the effect of these measures on the reproductive number.

Our data reveal that the seven different economic regions of Switzerland were sampled with different intensities. For this reason, we expect it is more reasonable to assume a homogeneous sampling intensity at the regional level than at the national level. On the other hand, infections imported from other regions likely represent a larger fraction of regional infections than imports from abroad do for national infections. However, the economic regions represent well-defined regions where we expect a lot of mixing within and less mixing across regions. Thus, we also performed analyses specific to the seven economic regions. Our regional estimates for B.1.1.7 transmission fitness advantage are largely in line with the national estimates, though of course with larger uncertainty. Two of the regions (Ticino, Central Switzerland) have too little data to make precise statements. Geneva is estimated to be around 2 weeks ahead in the B.1.1.7 dynamics compared to Switzerland as a whole. One explanation may be the large number of UK travelers to ski resorts in the Valais in December 2020.

Overall, we see a consistent signal for a large transmission advantage of B.1.1.7 in Switzerland across datasets and regions. Our results confirm other estimates from studies from the UK [1, 2, 3], Denmark [3], and Switzerland ([11]; looking at 501Y mutations). Furthermore, the strengthening of non-pharmaceutical interventions in Switzerland on 18 January 2021 could not stop the spread of B.1.1.7. We estimate that B.1.1.7 spread exponentially from its arrival in Switzerland in late 2020 until the end of our study period. Thus, despite the additional control efforts, total infections began growing exponentially again once B.1.1.7 became dominant in March 2021. Our core plots are available on [12] and [14] for real-time monitoring. Further, our model is integrated in the CoV-Spectrum website where it can be applied to other countries and variants [13].

## Supporting information

A7 D-BSSE GISAID Accession Numbers

A7 HUG GISAID Accession Numbers

## Data Availability

All used data and code are publicly available.

https://github.com/cevo-public/Quantification-of-the-spread-of-a-SARS-CoV-2-variant

## Contribution

CC: Conceptualization, Data curation, Formal Analysis, Investigation, Methodology, Resources, Software, Validation, Visualization, Writing - original draft; SN: Data curation, Writing - review & editing; IT: Data Curation, Resources, Software, Validation; MM: Formal Analysis, Methodology, Software, Writing - review & editing; JSH: Formal Analysis, Software, Visualization, Writing - review & editing; KPJ: Data curation, Formal analysis, Investigation, Methodology, Software, Validation, Visualization, Writing - review & editing; LF: Software; DD: Data curation, Software, Validation; KJ: Formal analysis, Validation; CA: Methodology, Writing - review & editing; NB: Conceptualization, Funding acquisition, Project administration, Supervision, Writing - review & editing; MA: Conceptualization, Investigation, Writing - review & editing; TS: Conceptualization, Funding acquisition, Investigation, Methodology, Project administration, Supervision, Validation, Writing - original draft; Everyone else: Resources, Writing - review & editing.

## Ethics

In this study we characterized the genome of viral RNA. As metadata, the date of sampling and location on a cantonal level was used. The local ethics commission (Ethikkommission Nordwest- und Zentralschweiz) confirmed that the study conforms to general ethical principal and that the study does not require ethical approval as only viral RNA with limited general metadata is used.

## Acknowledgement

We thank Richard Neher for useful discussions on the maximum likelihood inference. TS acknowledges funding from the Swiss National Science foundation (Special Call on Coronaviruses; 31CA30 196267 and 31CA30 196348). CA received funding from the European Union Horizon 2020 research and innovation programme - project EpiPose (No 101003688).

## A Supplementary Material

### A.1 Sequencing Protocols for Viollier samples and HUG samples

We perform whole-genome sequencing of the Viollier samples in three facilities. The HUG samples are processed in one of them, the Health 2030 Genome Center. The sequencing protocol for the samples sequenced at the Genomics Facility Basel and the Functional Genomics Center Zurich is described in [18]. Samples processed at the Health 2030 Genome Center used the Illumina COVIDSeq library preparation reagents following the protocol provided by the supplier [19]. These reagents are based on the ARTIC v3 multiplex PCR amplicon protocol [20]. When sufficient volume was available, 8.5ul of RNA extracted from patient nasopharyngeal samples were used in the cDNA synthesis step; if 8.5ul were not available, the maximum volume possible was used. Pooled libraries were sequenced on the Illumina NovaSeq 6000 using a 50-nucleotide pair-end run configuration. Post-sequencing library read de-multiplexing was done using an in-house developed processing pipeline [21]. The downstream bioinformatics procedure to obtain consensus sequences is described in [18, 22].

### A.2 Screening procedure at Dr Risch

Dr Risch medical laboratories used the Taqpath assay from Thermofisher for their diagnostic and recorded the S gene target failure (SGTF). SGTF samples are potential B.1.1.7 variants, as the B.1.1.7 variant causes a SGTF due to its deletion at positions 69-70 in the spike protein. Further, Dr Risch medical laboratories screen their samples for the 501Y mutation by a variant-specific PCR test. If a sample is identified as a potential VoC by these procedures, it was initially sent for whole-genome sequencing to the University Hospital Basel in order to confirm the B.1.1.7 variant. The sequencing protocol is described in [23]. For recent samples, the confirmation may still be outstanding or not conducted due to B.1.1.7 now being dominant. However, since typically a SGTF plus a 501Y mutation corresponds indeed to a B.1.1.7 variant, we consider these samples as B.1.1.7 variants even when whole genome sequencing confirmation is lacking.

### A.3 Estimating the transmission fitness advantage of a new variant

In what follows, we define two models describing the dynamics with which a new variant with a transmission fitness advantage spreads in a population. The first model is based on the assumption of discrete-time, while the second model is based on the assumption of continuous-time. Both models have been considered extensively in the literature to estimate fitness advantage (see e.g. [24, 3]). While in an epidemic, a generation does not end after a fixed time span (discrete-time model), generations are typically less variable than modeled under an exponential distribution (continuous-time model). Thus we view the two models as two extremes. We provide estimates based on both models and suggest that the true parameter may be anywhere within the ranges spanned by the two models. In the next sections, we provide details of how we estimate the transmission fitness advantage of B.1.1.7 based on daily data of the total number of samples and B.1.1.7 samples under these two models.

#### A.3.1 Discrete time model

We call *X* the common (non-B.1.1.7) variants and *Y* the B.1.1.7 variant. The process starts in generation 0 with *x*_0_ cases caused by variant *X* and *y*_0_ cases caused by variant *Y*. Let the number of cases in generation *n* be *x*(*n*) and *y*(*n*) for variants *X* and *Y*.

Let the reproductive number *R*_*d*_ of variant *X* in generation *n* be *R*_*d*_(*n*). Let the transmission advantage of variant *Y* be *f*_*d*_. Then the reproductive number of *Y* in generation *n* is (1 + *f*_*d*_)*R*_*d*_(*n*). Thus, we assume a multiplicative fitness advantage.

We have *x*(*n*) = *x*(0)×*R*_*d*_(0)*R*_*d*_(1) … *R*_*d*_(*n*−1) and *y*(*n*) = *y*(0)×*R*_*d*_(0)*R*_*d*_(1) … *R*_*d*_(*n*−1)(1+*f*_*d*_)^*n*^.

If *R*_*d*_ is constant through time, we have

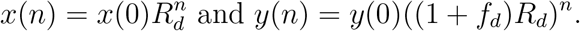

Let the proportion of variant *Y* at generation *n* be *p*(*n*). We have,

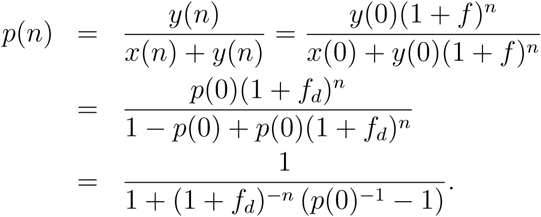

Thus, *p*(*n*) is the logistic function. It does not depend on *R*_*d*_.

If we write time in days *t* rather than generations *n* and assume a generation time of *g* days, we get

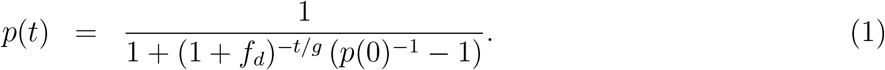

We now switch our parameterization to the more common

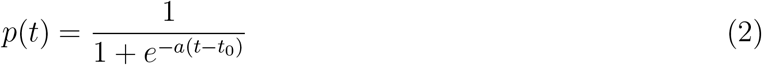

for parameter estimation from daily data. Parameter *a* is the logistic growth rate and parameter *t*_0_ the sigmoid’s midpoint.

The two free parameters, *a* and *t*_0_, are related to the two free parameters in Equation 1, *f*_*d*_ and *p*(0), as follows:

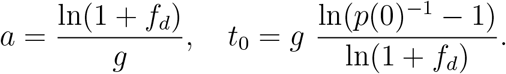

In particular, we get *f*_*d*_ = *e*^*ag*^ − 1.

#### A.3.2 Continuous time model

In continuous-time, instead of *R*_*d*_ and generation time *g*, we define the transmission rate *β* and the recovery rate *µ*. Under this model, the reproductive number is *R*_*c*_ = *β/µ*. Further, since an individual in the discrete model recovers after a generation of duration *g* (during which they left *R*_*d*_ offspring), we note that *g* is related to the expected time to recovery 1*/µ* in the continuous model, and in fact assume *g* = 1*/µ* in what follows. Again our initial numbers of the variants *X* and *Y* are *x*(0) and *y*(0). Calendar time is denoted by a continuous parameter *t*. We then have in expectation,

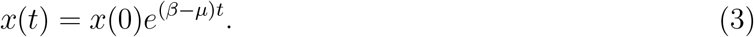

We note that *β* − *µ* is coined the Malthusian growth parameter [3].

Further, we again assume that variant *Y* has a transmission fitness advantage of *f*_*c*_, with transmission rate *β*(1 + *f*_*c*_) and recovery rate *µ*. The population size of the variant at time *t* is thus

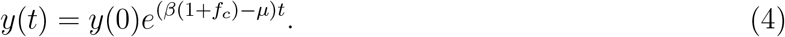

The proportion of the variant in the population at time *t* is

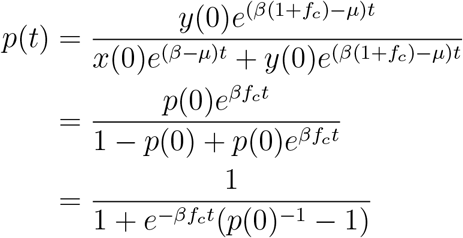

where we again recognize the logistic function. We turn again to the more common parameterization,

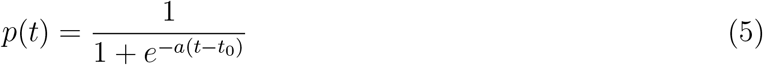

where we thus have 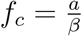.

The reproductive number is *R*_*c*_ = *β/µ* and the mean time to recovery, 1*/µ*, is equaled to *g*. Then, *β* = *R*_*c*_*/g*. Thus,

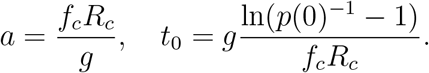

In particular, we have 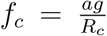. Note that the estimated fitness advantage under this model depends on the reproductive number *R*_*c*_ and is thus changing if *R*_*c*_ is changing through time.

#### A.3.3 Connection between discrete and continuous time

The discrete and continuous models are very similar. Both have the intitial conditions *x*(0) and *y*(0). For the dynamics, the discrete model has parameters *R*_*d*_ and *g* while the continuous model has parameters *β* and *µ*. We have *R*_*c*_ = *β/µ* and we further assumed that *g* = 1*/µ* (1*/µ* is the expected time until recovery in the continuous setting while *g* is the time to recovery in the discrete setting). The different parameterizations of fitness advantage are coined *f*_*c*_ and *f*_*d*_. We now determine how *f*_*c*_ and *f*_*d*_ are related.

To compare the two models, we now assume that their overall dynamics for the variant *X* are the same. After *n* generations of duration *g*, we have for variant *X*,

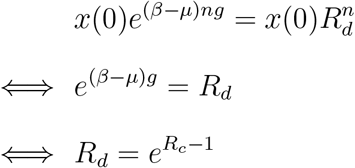

Using a Taylor expansion for *β/µ* close to 1, we obtain that indeed *R*_*d*_ = *R*_*c*_.

For the two models to produce the same growth also for variant *Y*, we require,

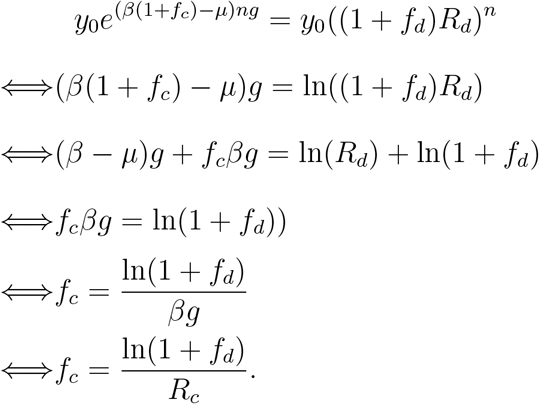

In the last step, we make use of *R*_*c*_ = *β/µ* and *g* = 1*/µ*.

This is equivalent to 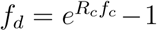. Using a Taylor expansion we get *f*_*d*_ = 1+*R*_*c*_*f*_*c*_ +*O*((*R*_*c*_*f*_*c*_)^2^)−1 and thus *f*_*d*_ = *R*_*c*_*f*_*c*_ for small *R*_*c*_*f*_*c*_.

#### A.3.4 Maximum likelihood parameter estimation

Next we explain how we estimate *a* and *t*_0_ of the logistic functions (Eqn. 2 and 5) from our data using maximum likelihood. We consider that we have data at times *t*_1_, …, *t*_*d*_. At time *t*_*i*_, we obtained *n*_*i*_ samples, where *n*_*i*_ is fixed, non-random.

We assume that the true number of B.1.1.7 variants at time *t*_*i*_ is a random variable, *K*_*i*_ which is binomially distributed with parameter *p*(*t*_*i*_), i.e.

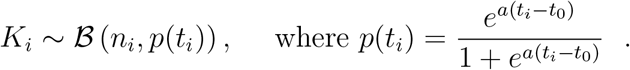

In particular, we assume here a deterministic logistic growth model for the increase in the proportion of variant *Y* (Eqn. 2 and 5), on top of which only the drawing process is random. This model simplifies naturally to a very popular logistic regression. This is an instance of a Generalized Linear Model, where the natural parameter of the binomial distribution is a linear function of predictors, the only predictor considered here being the time *t*.

We use the *Python* library *statsmodel* [25] to recover maximum likelihood estimates (MLEs) and confidence intervals. The confidence intervals are based on an asymptotic Gaussian distribution for the parameters of the logistic regression fitted to our data, i.e. the fixed values *t*_1_, …, *t*_*d*_, *n*_1_, …, *n*_*d*_ as well as the numbers of samples at each time point being the variant B.1.1.7, *k*_1_, …, *k*_*d*_. Given the large number of sequences – we have at least 100 samples per region – the use of an asymptotic Gaussian approximation is justified. Parameters *a, t*_0_, *f*_*d*_, *f*_*c*_ as well as the proportions of variant B.1.1.7 *p*(*t*) through time are simple transformations of the parameters of the logistic regression. Their MLEs are the same transformations applied to the MLEs of the logistic regression parameters. The difference between the MLEs and the true parameters are again Gaussian, with a covariance matrix found by applying the delta method. This is used to construct confidence intervals for all these quantities. We used the default fitting procedure provided in the *statsmodel* package. This procedure reported convergence for all analyses.

### A.4 Estimation of the effective reproductive number

We use the number of confirmed cases per day from the Federal Office of Public Health, Switzerland, for 14 December 2020 to 11 March 2021. Then, for each day, we estimate the number of B.1.1.7 variants by multiplying the total number of confirmed cases by the proportion of B.1.1.7 in our dataset (Viollier or Risch). We then estimate an effective reproductive number of the B.1.1.7 variant and of the non-B.1.1.7 variants using these data. For this estimation, we use the method developed in [26]. This method consists of two main parts: first, the observed case data is related to the corresponding time series of infections. We smooth the observations using LOESS smoothing to remove weekend effects. Then, we deconvolve with the delay of infection to symptom onset (gamma distributed with mean 5.3 and sd 3.2) and the delay from symptom onset to case confirmation (gamma distributed with mean 5.5 and sd 3.8). Second, we estimate the effective reproductive number from the time series of infection incidence using EpiEstim [27]. The reported point estimate is the estimate on the original case data. To account for uncertainty in the observation process, the observed daily case incidences are additionally bootstrapped 1000 times, resulting in an ensemble of alternative case incidence time series and corresponding estimated effective reproductive numbers. These are used to construct the 95% confidence interval around the effective reproductive number, and to calculate the standard deviation of the ratios of effective reproductive number estimates (see below).

We perform the estimation of the reproductive number in two different ways. First, we estimate smooth changes in the reproductive number, by estimating it across the entire time series using a 3-day sliding window. Second, we assume the reproductive number was constant during time intervals in which the non-pharmaceutical interventions did not change. Since 18 January 2021, Switzerland has implemented a set of tighter measures (in particular, shops are closed and the size of gatherings is restricted to five people [28]). Thus we fix the reproductive number to be constant between 01 January and 17 January 2021. Then the reproductive number is allowed to change and again fixed to be constant from 18 January 2021 onwards.

To compare the effective reproductive number *R* of the B.1.1.7 variant (*Y*) to that of non-B.1.1.7 variants (*X*), we take the ratio 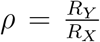 at every time point. The standard deviation of this ratio *σ*_*ρ*_ was found through Gaussian error propagation of the standard deviation of the individual *R* estimates (*σ*_*X*_, *σ*_*Y*_):

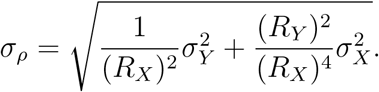

**Figure S1:**
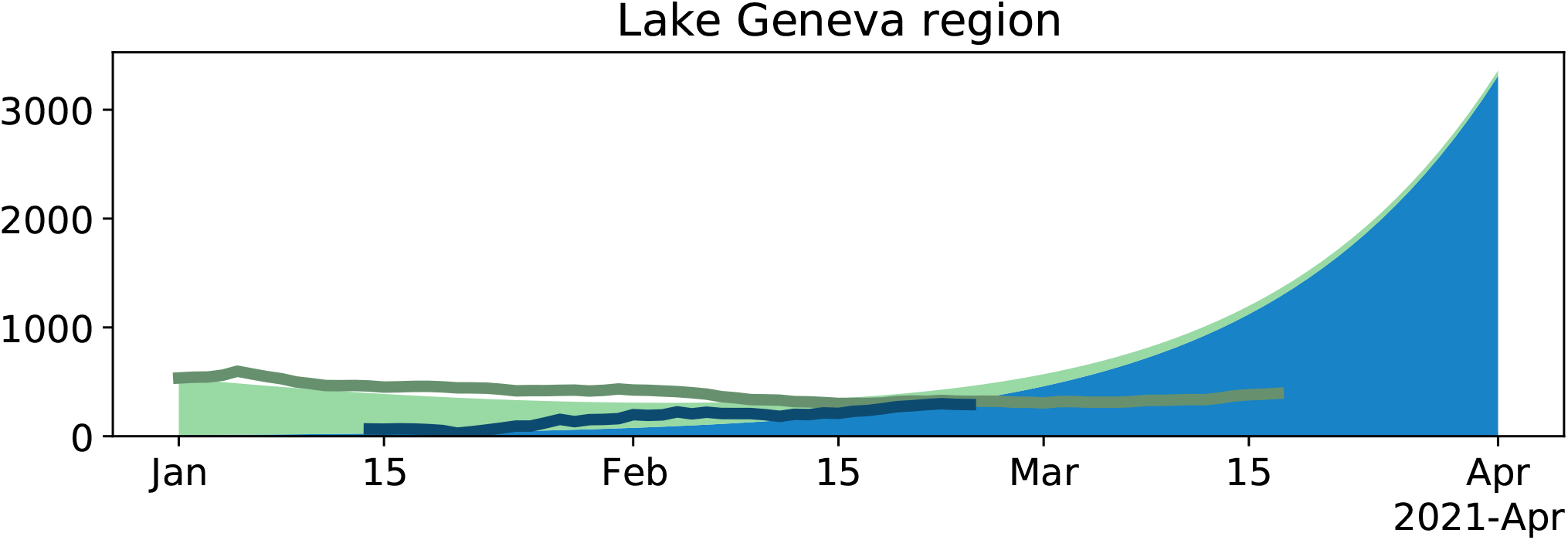
Change in the number of B.1.1.7 variants and in the number of all cases through time for the Lake Geneva region. For details see legend of Fig. 4. Compared to 4, we here use the average reproductive number estimated for non-B.1.1.7 in Geneva for the time period 01 January 2021-17 January 2021. The transmission fitness advantage is calculated based on this reproductive number and the estimate of the growth rate *a* for the Lake Geneva region.

### A.5 Discrepancy for Ticino and Lake Geneva

The discrepancy for Ticino and Lake Geneva is not surprising: they had a reproductive number which was different from the national reproductive number in the first half of January. For Ticino, the empirical case numbers drop faster than the model, which is in line with a lower reported reproductive number compared to the national level[17]. For the Lake Geneva region, the empirical case numbers drop slower than the model, which is in line with a higher reported reproductive number compared to the national level[17]. For all regions but Ticino, we have enough data to estimate a reproductive number for the non-B.1.1.7 variants for 01 January-17 January 2021. While for Switzerland, we obtained a point estimate of 0.83, the point estimates for all regions but Lake Geneva are between 0.81-0.83. Thus using the point estimate for all of Switzerland for the regional plots - with the exception of Lake Geneva and Ticino - in Fig. 5 is justified. For Geneva, we obtain a point estimate of 0.88. We use this point estimate in a Lake Geneva specific model (Fig. S1). Again we observe that the total number of confirmed cases dropped recently faster compared to the model. For a discussion on the discrepancy see main text.

**Figure S2:**
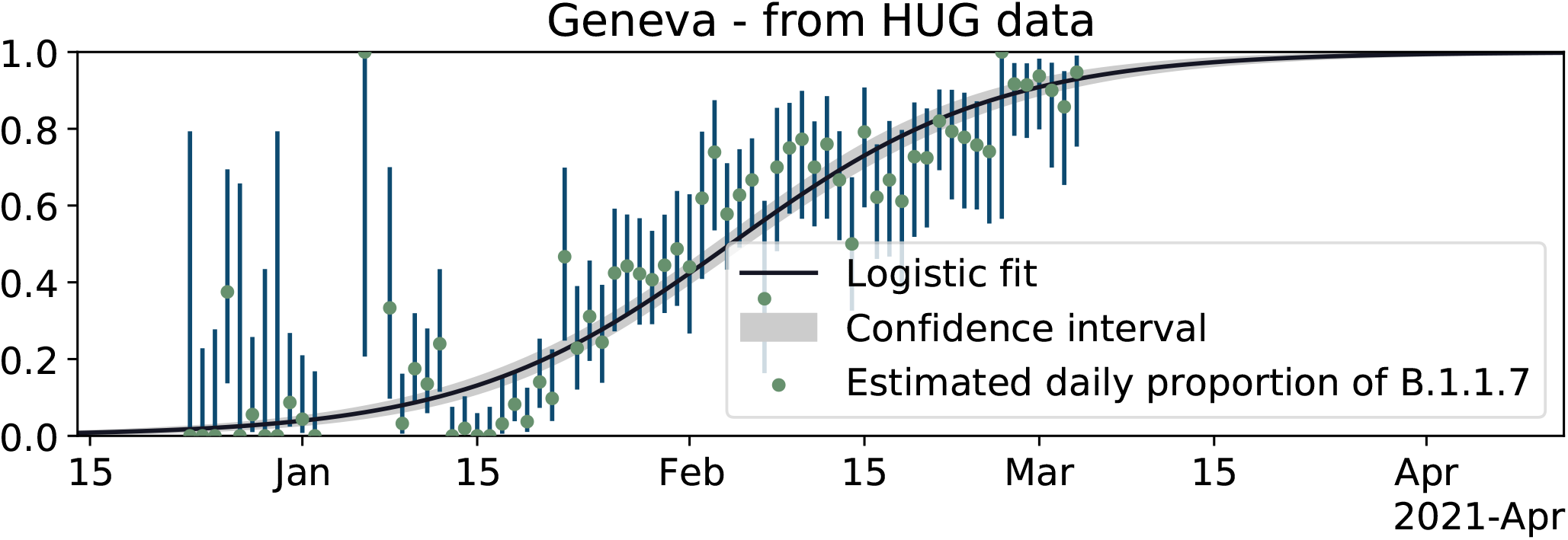
Logistic growth of frequency of B.1.1.7 in the Lake Geneva region based on HUG data.

### A.6 Additional Tables

**Table S1:**
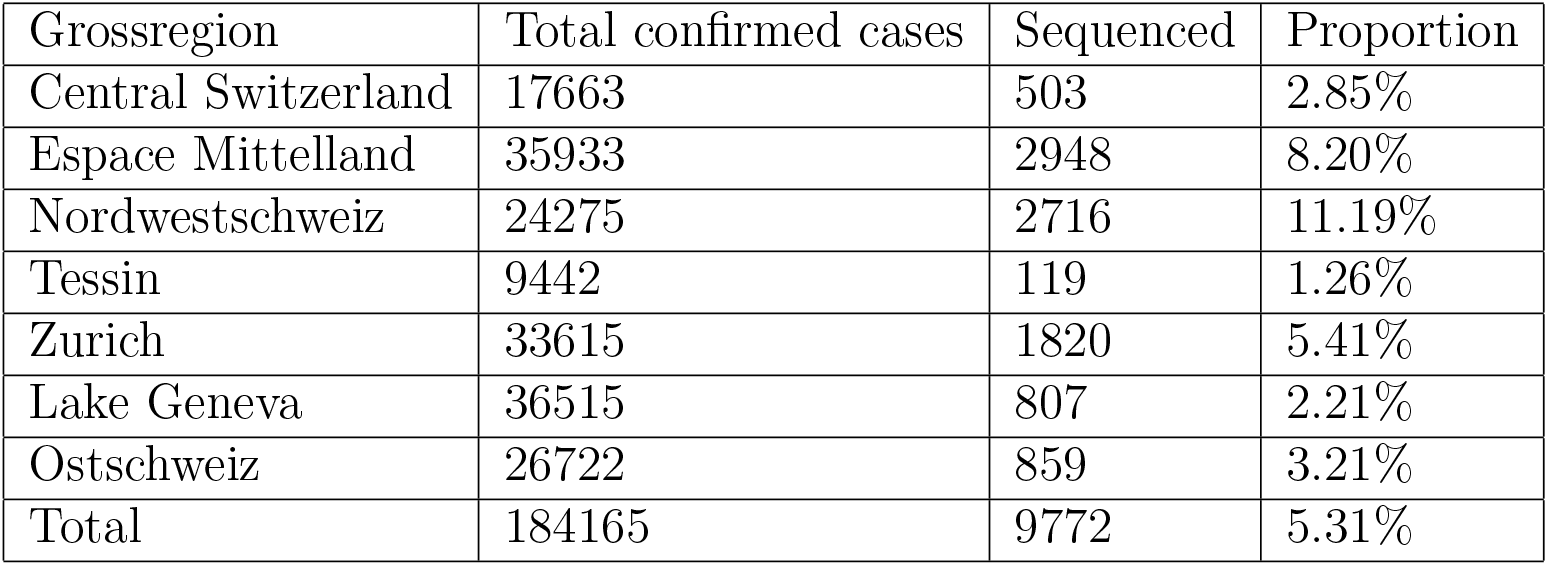
The proportion of sequenced cases out of all cases for the Viollier dataset.

**Table S2:** The proportion of characterized cases out of all cases for the Risch dataset.

| Grossregion | Total confirmed cases | Sequenced | Proportion |
| --- | --- | --- | --- |
| Central Switzerland | 17663 | 321 | 1.82% |
| Espace Mittelland | 35933 | 3893 | 10.83% |
| Nordwestschweiz | 24275 | 1135 | 4.68% |
| Tessin | 9442 | 113 | 1.20% |
| Zurich | 33615 | 1090 | 3.24% |
| Lake Geneva | 36515 | 405 | 1.11% |
| Ostschweiz | 26722 | 4594 | 17.19% |
| Total | 184165 | 11551 | 6.27% |

### A.7 GISAID Accession Numbers

The used sequences were, if fulfilling the quality criteria, uploaded to GISAID. The GISAID accession numbers are in the files supplementary material a7 d-bsse gisaid ids.txt and supplementary material a7 hug gisaid ids.txt which are attached to this paper.

